# Monkeypox virus contamination in an office-based workplace environment, England 2022

**DOI:** 10.1101/2022.08.09.22278460

**Authors:** Barry Atkinson, Susan Gould, Antony Spencer, Okechukwu Onianwa, Jenna Furneaux, James Grieves, Sian Summers, Tim Crocker-Buqué, Tom Fletcher, Allan M Bennett, Jake Dunning

**Author notes:** Correspondence: Barry Atkinson. These authors contributed equally to this work and share first authorship (Barry Atkinson, Susan Gould).

## Abstract

In May 2022, an office worker attended their place of work while experiencing prodromal symptoms of monkeypox infection. Environmental sampling performed four days later identified only low levels of monkeypox virus DNA contamination of the worker’s desk, and no contamination elsewhere within the office. Replication-competent virus was not identified.

## [Untitled Introduction]

More than 16,000 cases of monkeypox have been reported globally in 2022, predominately in non-endemic countries (1). Although transmission in the current outbreak is typically via prolonged direct contact with confirmed cases (2), infection-competent monkeypox virus (MPXV) has been recovered from contaminated environments multiple days after last occupancy (3) raising the potential for fomite transmission. In addition, prolonged close contact such as working in an open-plan office could result in respiratory droplet transmission of MPXV (4,5).

### Sampling Location

In May 2022, an individual working in a non-clinical role in an administrative office within a hospital acquired MPXV infection following non-occupational exposure. The individual worked in a 15-desk open-plan office for one working day following onset of a mild, influenza-like illness, and took steps to reduce mixing and avoid close contact with others. Several COVID-19 control measures were still implemented within this office including a requirement to wear medical masks and regular hand hygiene. In addition, this office had permanent partitions between desk spaces, approximately 1.2 metres high. The individual reported that monkeypox skin lesions appeared two days after taking sickness absence and at that point the office was closed to all staff, pending a risk assessment and risk management plan. 17 staff contacts were identified, including six category 2 and four category 1 contacts according to UKHSA contact categorisation (6); four of these individuals accepted post-exposure prophylaxis with Imvanex^®^ vaccine when it was offered according to UKHSA guidelines. No contacts developed symptoms consistent with monkeypox during their 21-day monitoring periods.

A decision was made to perform cleaning and decontamination of the office given its location within a healthcare facility and due to the environmental stability of orthopox viruses. This was performed by professional decontamination staff following a protocol used during previous monkeypox outbreaks (7). All papers, disposable items, and anything that could not be decontaminated per protocol were removed as waste. Surface activator was applied to all hard surfaces followed by 1000ppm hypochlorite solution that was allowed to air dry. Soft surfaces were decontaminated by mechanical vacuuming with HEPA filtration, followed by steam cleaning. The hospital performed a final decontamination of the office using hydrogen peroxide vapour (Bioquell BQ-50 with 35% hydrogen peroxide solution).

### Environmental Sampling

Prior to decontamination, environmental sampling was performed to identify MPXV contamination. Sampling was performed four days after the case was last in the office and two days after the office was closed to all staff. Surface samples were collected from non-porous surfaces such as desks and telephones using Copan UTM^®^ swabs, and from porous surfaces such as carpets and chair seats using the Sartorius MD8 Airport with gelatine filters. In addition, SKC wearable samplers were utilised during the sample collection process to measure any re-aerosolisation of MPXV. All samples were processed as previously described (8) and analysed for the presence of MPXV DNA using rRT-PCR as previously reported (3,9).

### Monkeypox Virus Contamination

Only 3/34 surface samples were positive for the presence of MPXV DNA with all positive samples returning crossing threshold (Ct) values above 34 cycles indicating low-level contamination (Figure 1). All three positive samples were from the case’s desk area including their telephone (Ct 37.7), keyboard (Ct 36.9) and a 10×10cm area of their desk (Ct 34.3). Five other surface samples collected from the case’s desk were negative for MPXV DNA including chair armrest, desk partition and computer mouse, as were 26 surface samples collected from other desks and high-touch areas throughout the office. All non-porous samples were negative for MPXV DNA, as were both wearable air samples.

**Figure 1:**
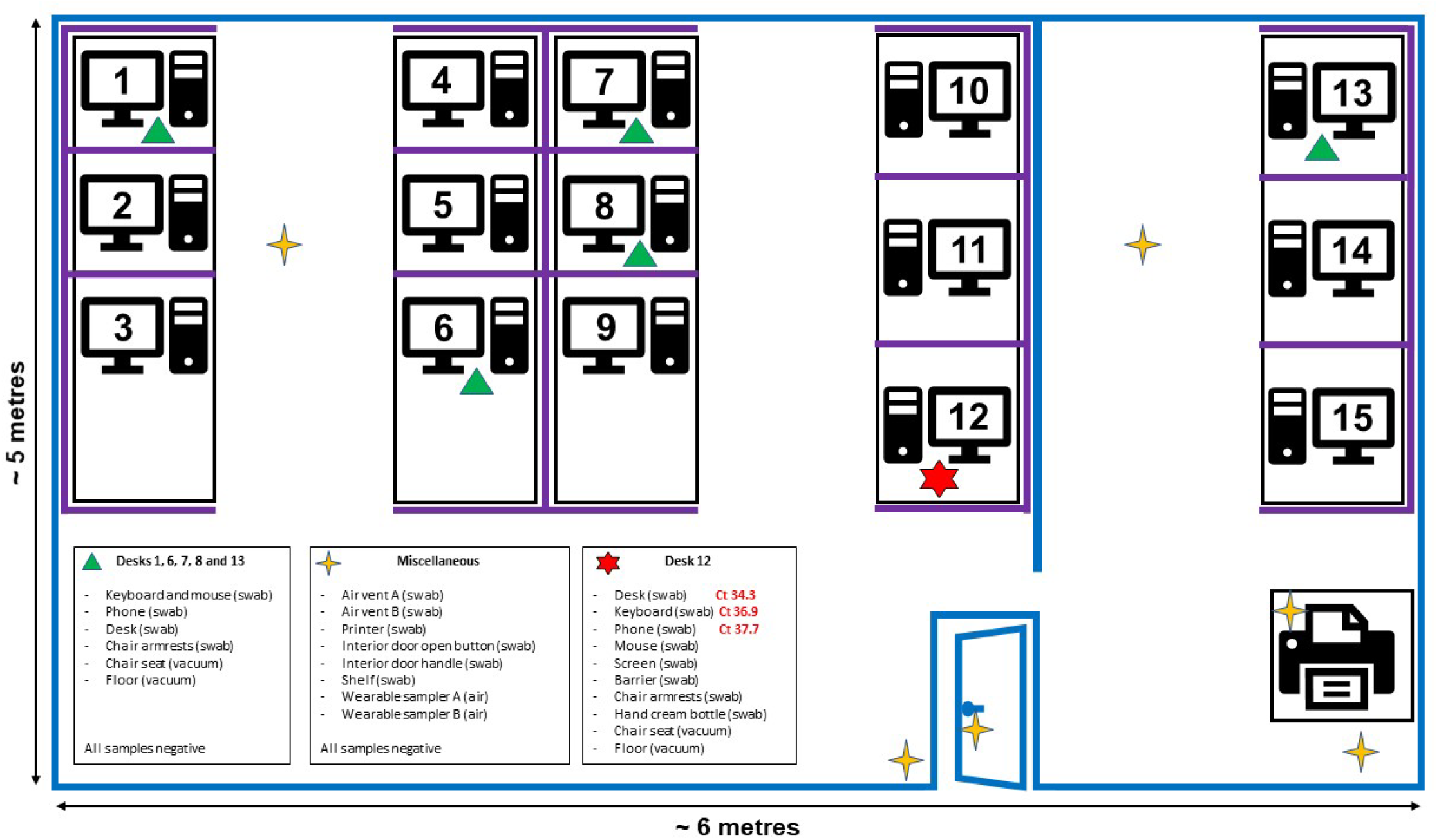
Diagrammatic representation of the office environment associated with a confirmed case of monkeypox. Blue lines represent permanent office structures such as walls and office door; purple lines represent desk partitions (wooden partitions approximately 1.2 metres high enclosing work desks). Ct = crossing threshold value of MPXV DNA detected in sample.

Virus isolation was attempted on the Ct 34.3 positive desk sample using a previously described method (8); no evidence of replicating virus or cytopathic effect was observed after 10 days of monitoring.

## Conclusions

Environmental sampling performed in an open-plan office that had been occupied by a person with monkeypox during their first 24 hours of illness identified low-level MPXV DNA contamination localised to their immediate desk area. Attempts to isolate virus from the most positive sample (Ct 34.4) were unsuccessful, suggesting the absence of replication-competent virus. As sampling was performed four days after occupancy by the infected individual, it is possible that some level of DNA or viral degradation occurred prior to sampling, although the office was windowless (minimising UV light degradation), the office was not cleaned prior to sampling, and MPXV is known to be environmentally stable.

It is notable that the patient reported their skin lesions only emerged after they had taken leave from work due to illness, raising the possibility that the MPXV DNA detected on their desk may have come from respiratory secretions through droplets or contaminated hands. If so, and while we do not know the amount of time that the individual’s medical mask remained in place, it is possible that the use of the medical mask may have reduced environmental contamination by respiratory droplets containing virus. Unfortunately, the individual’s disposable medical mask was not retained and therefore could not be tested for the presence of MPXV DNA.

Although this office may be similar to other offices in design, our findings should be seen as context-specific, including the fact that the individual worked only during the early ‘prodromal’ phase of their monkeypox illness, several COVID-19 measures were still in place including a requirement to wear medical masks, and physical partitions were present between desk spaces. The limited detection of MPXV DNA and absence of secondary cases do not demonstrate that cleaning is unnecessary in an office where an infected person has worked, or that focussed cleaning of an infected person’s desk area is all that is required. In the absence of reliable, real-time environmental sampling to inform decontamination, and the fact that the office was within a hospital, our detection of environmental MPXV DNA supports the decision made to remediate the entire office. These data confirm that MPXV contamination can occur in workplace environments occupied by a person with early monkeypox illness and, accordingly, appropriate cleaning and decontamination measures should be considered in such situations.

## Data Availability

All data produced in the present work are contained in the manuscript

## Statements

### Funding

This work was funded by UKHSA Grant in Aid funding and the NIHR Health Protection Research Unit in Emerging and Zoonotic Infections. The funding source had no involvement in study design; in the collection, analysis and interpretation of data; in the writing of the report; or in the decision to submit the article for publication. JD is supported by the Moh Foundation.

### Competing interests

None declared.

### Ethical approval

The investigations performed were a component of the urgent public health investigation performed as part of UKHSA’s public health incident response to cases of a high consequence infectious disease in the UK. UKHSA is the national health security agency for England and an executive agency of the UK Government’s Department of Health and Social Care. The study protocol was subject to internal review by the Research Ethics and Governance Group, which is the UKHSA Research Ethics Committee, and was granted full approval.

## Acknowledgments

The authors wish to acknowledge Ambipar Response Ltd for providing information on their decontamination process.

## Authors’ contributions

Conceptualisation and methodology: BA, SG, TF, AMB and JD.

Investigation: BA, SG, T-CB and JD.

Formal analysis: BA, AS, OO, JF, JG and SS.

Writing – original draft: BA, SG, TF, AMB and JD.

Writing – review and editing: All authors.

## Disclosure Statement

This report contains work supported by UKHSA Grant-in-Aid. The contents of this paper, including any opinions and/or conclusions expressed, are those of the authors alone and do not necessarily reflect UK Health Security Agency policy.

## Notes

### Competing Interest Statement

The authors have declared no competing interest.

### Author Declarations

The investigations performed were a component of the urgent public health investigation performed as part of the UKHSA public health incident response to cases of a high consequence infectious disease in the UK. UKHSA is the national health security agency for England and an executive agency of the UK Government Department of Health and Social Care. The study protocol was subject to internal review by the Research Ethics and Governance Group, which is the UKHSA Research Ethics Committee, and was granted full approval.

## References

1. WHO. Multi-country outbreak of monkeypox, External situation report #2 - 25 July 2022 [Internet]. 2022 [cited 2022 Jul 26]. Available from: https://www.who.int/publications/m/item/multi-country-outbreak-of-monkeypox--external-situation-report--2---25-july-2022

2. Cohen. Monkeypox outbreak questions intensify as cases soar | Science | AAAS [Internet]. 2022 [cited 2022 Jun 1]. Available from: https://www.science.org/content/article/monkeypox-outbreak-questions-intensify-cases-soar

3. Atkinson B, Burton C, Pottage T, Thompson K-A, Ngabo D, Crook A, et al. Infection-competent monkeypox virus contamination identified in domestic settings following an imported case of monkeypox into the UK. Environ Microbiol. 2022 Jul 15;

4. Ježek Z, Grab B, Szczeniowski MV, Paluku KM, Mutombo M. Human monkeypox: secondary attack rates. Bull World Health Organ. 1988;66(4):465–70.

5. Hutson CL, Carroll DS, Gallardo-Romero N, Weiss S, Clemmons C, Hughes CM, et al. Monkeypox disease transmission in an experimental setting: prairie dog animal model. PloS One. 2011;6(12):e28295.

6. UKHSA. Monkeypox: contact tracing [Internet]. GOV.UK. [cited 2022 Jul 26]. Available from: https://www.gov.uk/government/publications/monkeypox-contact-tracing

7. Public Health England. Monkeypox: Guidance for environmental cleaning and decontamination - version 4. https://assets.publishing.service.gov.uk/government/uploads/system/uploads/attachment_data/file/746086/Monkeypox_Guidance__cleaning_decontamination.pdf [Internet]. Public Health England; 2018. Available from: https://assets.publishing.service.gov.uk/government/uploads/system/uploads/attachment_data/file/746086/Monkeypox_Guidance__cleaning_decontamination.pdf

8. Gould S, Atkinson B, Onianwa O, Spencer A, Furneaux J, Grieves J, et al. Air and surface sampling for monkeypox virus in UK hospitals. medRxiv. 2022 Jul 21;2022.07.21.22277864.

9. Li Y, Zhao H, Wilkins K, Hughes C, Damon IK. Real-time PCR assays for the specific detection of monkeypox virus West African and Congo Basin strain DNA. J Virol Methods. 2010 Oct;169(1):223–7.

